# Behavioral preventive measures and the use of medicines and herbal products among the public in response to Covid-19 in Bangladesh: A cross-sectional study

**DOI:** 10.1101/2020.08.15.20175513

**Authors:** Iftekhar Ahmed, Maruf Hasan, Rahima Akter, Bidduth Kumar Sarkar, Marufa Rahman, Md Samun Sarker, Mohammed A. Samad

**Affiliations:** Department of Pharmacy, Jahangirnagar University, Savar, Dhaka, Bangladesh; Department of Pharmacy, World University of Bangladesh, Dhaka, Bangladesh; Department of Pharmacy, Ranada Prasad Shaha University, Narayanganj, Bangladesh; Bangladesh Livestock Research Institute (BLRI), Savar, Dhaka, Bangladesh

**Keywords:** Covid-19, Preventive measures, Medication use, Herbal products use, Bangladesh

## Abstract

The present study was conducted to assess the behavioral preventive measures and the use of medicines and herbal foods/products among the public in response to Covid-19. A cross-sectional survey was conducted from 27 June to 20 July 2020, and 1222 people participated. Kruskal-Wallis test was used to identify the differences in behavioral preventive practices across different demographic categories. To identify the factors associated with the use of preventive medicines and herbal foods/products, multivariable logistic regression was performed. Most participants adopted the recommended preventive practices such as washing hands more frequently (87.5%), staying home more often (85.5%), avoiding crowds (86%), and wearing masks (91.6%). About half of the smokers reported a decreased rate of smoking during the pandemic. Also, 14.8% and 57.6% of the participants took medicines and herbal foods/products as preventive measures against Covid-19. Arsenicum album and Zinc supplements were the most commonly used preventive medicines. Gender, age, and fear of Covid-19 were significantly associated with the use of both preventive medicines and herbal products. For the management of Covid-19 related symptoms, Paracetamols, Fexofenadine, and Zinc supplements were used most often. Most participants sought information from non-medical sources while using medicines and herbal products. Moreover, potentially inappropriate and unnecessary use of drugs were identified.

## Introduction

Bangladesh is one of the most affected countries by the Covid-19 pandemic, with 232,194 confirmed cases as of 30 July 2020 [1]. As a resource-limited country, Bangladesh is already dealing with a multitude of challenges pertaining to the pandemic starting from insufficient testing, lack of public awareness, and inadequate facilities to treat patients who are infected with Severe acute respiratory syndrome coronavirus 2 (SARS-CoV-2) [2, 3]. For instance, the capability of the government to treat infected patients is extremely limited, with only 733 intensive care unit (ICU) beds and fewer than 1,800 ventilators for critical support [4]. Moreover, there have been reports of patients being denied treatments from public and private hospitals, mostly because most hospitals do not have the equipment to treat patients, which is also causing unwanted deaths [4, 5].

Given this situation, hospital-based interventions cannot be a primary means of dealing with the pandemic in resource-limited countries like Bangladesh; instead, efforts must be taken to prevent the spread of the virus as much as possible. Recommended preventive measures to stifle the spread of Covid-19 include practices such as wearing masks, avoiding crowds, washing hands regularly, staying home as much as possible, etc. [6]. Furthermore, in absence of any specific vaccines or antiviral therapy against Covid-19, experts have recommended the intake of vitamins, minerals, and herbal medicines in order to bolster the immune system which can lower the risk and severity of infection [7–9].

The panic and fear surrounding the pandemic combined with misinformation have resulted in panic buying and hoarding of medicines by the public [10, 11]. Also, because of the lockdown, access to health providers have been restricted significantly, and that is likely to make people more prone to self-medication and more dependent on less reliable sources such as the social and digital media for medicine-related information. Therefore, it is very likely that there have been inappropriate and unnecessary uses of medicines by the public in relation to the prevention and cure of Covid-19 symptoms.

The primary focus of this study was to explore what medicines and herbal products/foods are being used by the public as preventive measures against Covid-19, as well as to manage symptoms related to Covid-19. To our best knowledge, this is the first study in any country to assess the use of medications among the public in response to Covid-19. We also explored the behavioral preventive measures among the public, rather briefly, since a number of studies have already been conducted to assess preventive practices of the public in relation to Covid-19 in Bangladesh [12–14].

## Methods

### Study design and sampling

We performed a cross-sectional survey from 27 June to 20 July 2020 using a self-administered questionnaire. Data collection was conducted both in-person and online. The online survey was conducted using google form and participants were recruited via social media platforms such as Facebook and WhatsApp. The in-person survey was done in different parts of the country including Dhaka, Manikganj, Cumilla, Tangail, and Nilphamari. Adults age 18 years and above were eligible to participate in the survey. The research protocol was reviewed and approved by the Antimicrobial Resistance Action Center (ARAC), Animal Health Research Division, Bangladesh Livestock Research Institute (BLRI), Bangladesh (Approval no: ARAC:15/06/2020:03). Participants were made aware of the purpose of the study and informed consent was taken from them.

### The questionnaire

The questionnaire comprised of three parts. The first part was related to sociodemographic information such as age, gender, education, marital status, place of residence (rural or urban), presence of chronic disease, as well as participants’ fear of Covid-19. The second part contained four questions related to behavioral preventive measures adopted by the people such as washing hands, staying home, avoiding crowds, and wearing masks. Participants were also asked if there had been any changes in their smoking habits during the pandemic.

The third section asked questions about the participants’ use of medicines and herbal products/foods as preventive and curative measures against Covid-19. First, participants were asked whether they had experienced any Covid-19 related symptom or not. Those who did not experience any symptoms were asked if they had taken any medicine or herbal product as a preventive measure to lower the risk of infection. As for those who experienced one or more symptoms related to Covid-19, they were asked what medicines they had used to manage those symptoms, as well as if they had taken any medicine or herbal products/foods as a preventive measure prior to the occurrence of symptoms. For this purpose, a list of the common symptoms of Covid-19 (fever, dry cough, tiredness, sore throat, difficulty breathing) based on the WHO guidelines was provided. Here it should be mentioned that we relied on participants’ self-reports of symptoms instead of Covid-19 test results since only a very small percentage of the population is being tested, with only 6,985 people per million population [1]. Participants were also asked about the source of information/advice regarding their medication and herbal product usage. The questionnaire was translated to Bengali and pretested in a pilot survey of 10 people, and amendments were made where necessary.

### Statistical analysis

The sociodemographic characteristics of the participants, the frequency of behavioral preventive practices, the use of medicines and herbs, as well as the source of medication-related information were reported using descriptive statistics. The number of behavioral preventive practices (washing hands, staying home, avoiding crowds, and wearing masks) adopted by each participant was calculated and was expressed as continuous data ranging from 0 to 4. To identify the differences in behavioral preventive practices across different demographic categories, the Kruskal-Wallis nonparametric test was performed. The Kruskal-Wallis test was preferred over ANOVA because the data was not normally distributed. To assess the factors associated with the use of preventive medicines and herbal foods/products, multivariable binary logistic regression was performed, calculating the adjusted odds ratios (OR) with 95% confidence interval (CI). A *P*-value of less than 0.05 was considered statistically significant. Data analysis was performed in IBM SPSS version 25.

## Results

### Participants’ characteristics

A total of 1222 people participated in the survey. The age of the participants ranged from 18 to 82 years, and the mean age was 30.77 (SD: 12.1 years). Among the respondents, 61.4% were male, 61.9% lived in urban areas, and 52.1% had received university (undergraduate and graduate) education. The fear of Covid-19 varied, with 15.6% reporting being not afraid at all while 28 % said they were very afraid. Detailed characteristics of the participants are provided in table 1.

**Table 1:** Sociodemographic Characteristics of the participants

| **Characteristic** | **Frequency** | **Percentage** |
| --- | --- | --- |
| **Gender** |  |  |
| Male | 750 | 61.4 |
| Female | 466 | 38.1 |
| **Age** |  |  |
| 18-29 | 767 | 62.8 |
| 30-44 | 277 | 22.7 |
| 45-59 | 112 | 9.2 |
| 60 and greater | 60 | 4.9 |
| **Residence** |  |  |
| Rural | 461 | 37.7 |
| Urban | 756 | 61.9 |
| **Education** |  |  |
| Secondary or lower | 334 | 27.3 |
| Higher secondary | 245 | 20.0 |
| Undergraduate degree | 410 | 33.6 |
| Graduate degree | 226 | 18.5 |
| **Marital status** |  |  |
| Unmarried | 662 | 54.2 |
| Married | 559 | 45.7 |
| **Presence of chronic disease** |  |  |
| No | 1052 | 86.1 |
| Yes | 166 | 13.6 |
| **Fear of covid-19** |  |  |
| Not afraid | 191 | 15.6 |
| Somewhat/ Moderately afraid | 683 | 55.9 |
| Very afraid | 342 | 28.0 |

### Behavioral preventive measures

Most of the participants reported following the behavioral preventive guidelines such as frequent washing of hands (87.5%), staying home more often (85.5%), avoiding crowds (86%), and wearing masks (91.6%) (figure 1a). Compliance with these practices was significantly higher among the highly educated, females, younger participants, participants living in urban areas, and those with a higher level of Covid-19 fear (table 2). Moreover, 25.6% (313) identified themselves as smokers, and 48.6% (152) of them said they were smoking less frequently than before the pandemic (figure 1b).

**Figure 1:**
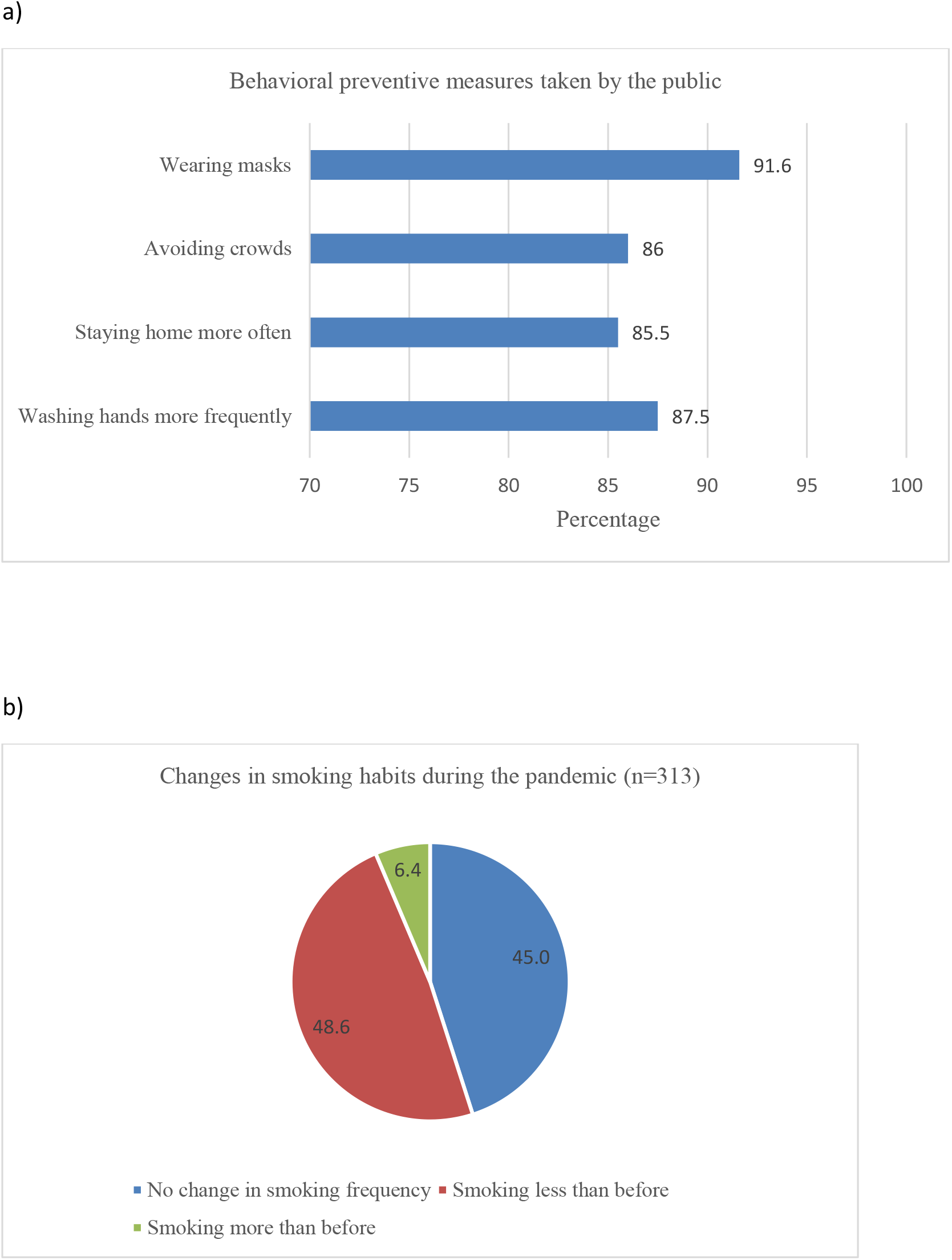
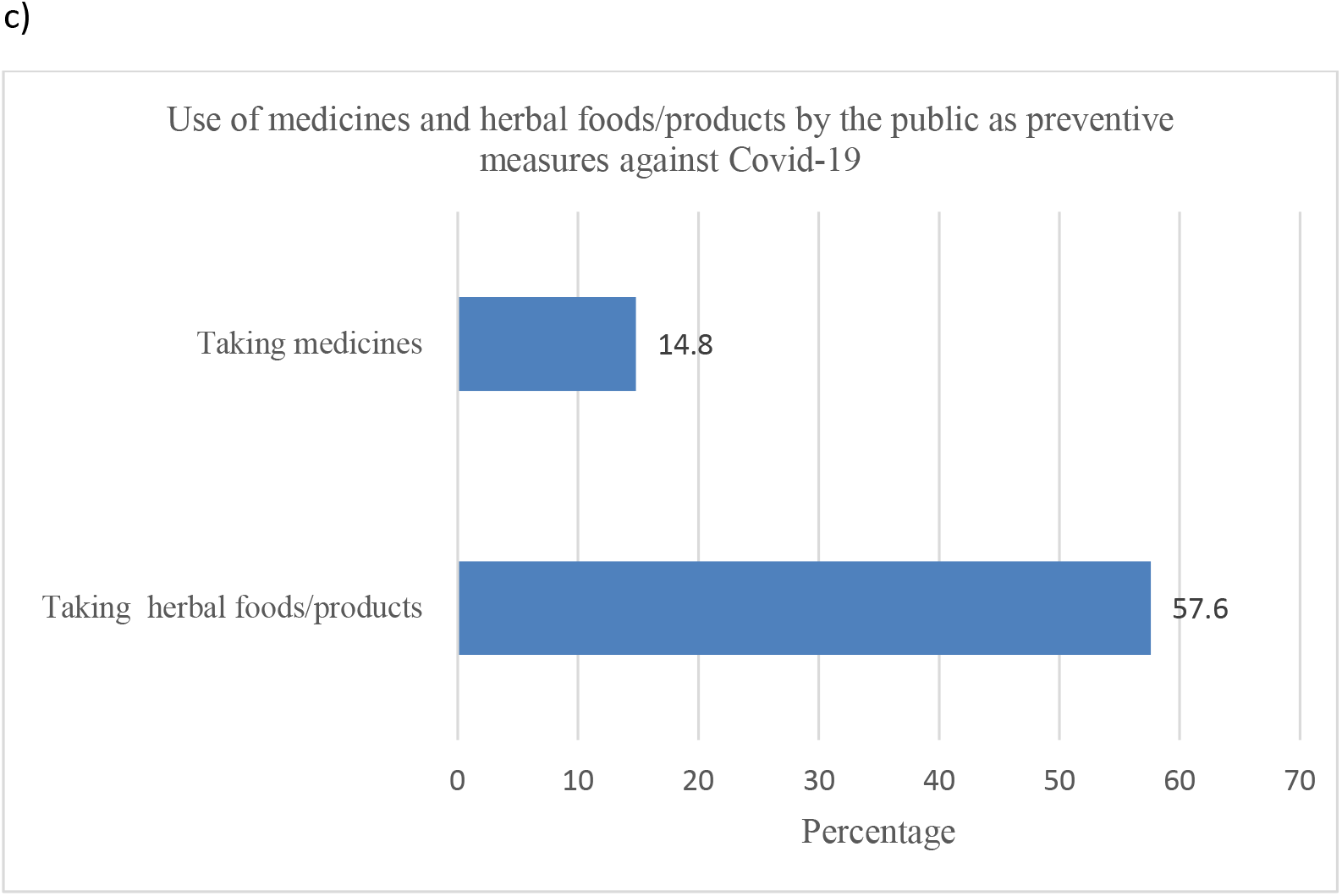
Behavioral preventive measures (a, b) and the use of preventive medicines and herbal products (c) among the public in response to Covid-19

**Table 2:** Groupwise means and the results of the Kruskal-Wallis test on the differences in the number of behavioral preventive practices across different demographic categories.

| **Variables** | **Number of preventive practices (Mean)** | **Standard Deviation** | **P-value** |
| --- | --- | --- | --- |
| **Gender** |  |  | <0.001 |
| Male | 3.35 | 1.09 |  |
| Female | 3.76 | 0.58 |  |
| **Age** |  |  | 0.049 |
| 18-29 | 3.54 | 0.93 |  |
| 30-44 | 3.48 | 0.95 |  |
| 45-59 | 3.37 | 0.1 |  |
| 60 and greater | 3.37 | 1.06 |  |
| **Residence** |  |  | <0.001 |
| Rural | 3.25 | 1.17 |  |
| Urban | 3.66 | 0.74 |  |
| **Education** |  |  | <0.001 |
| Secondary or lower | 3.00 | 1.25 |  |
| Higher secondary | 3.62 | 0.85 |  |
| Undergraduate | 3.71 | 0.71 |  |
| Graduate | 3.76 | 0.56 |  |
| **Marital status** |  |  | 0.005 |
| Unmarried | 3.57 | 0.89 |  |
| Married | 3.43 | 1.0 |  |
| **Presence of chronic disease** |  |  | 0.81 |
| No | 3.50 | 0.96 |  |
| Yes | 3.52 | 0.87 |  |
| **Fear of covid-19** |  |  | <0.001 |
| Not afraid | 2.76 | 1.42 |  |
| Somewhat/ Moderately afraid | 3.57 | 0.81 |  |
| Very afraid | 3.80 | 0.6 |  |

### Medication use

About 15% (181) of the participants took conventional and homeopathic medicines prophylactically in order to lower the risk of being infected with the coronavirus. The most commonly used medicine was Arsenicum album, a homeopathic formulation of Arsenic trioxide. Other commonly used drugs were Zinc supplements, Vitamins, and Paracetamols (figure 2a). Results of regression analysis show that males were more likely to take preventive medicines compared to females (OR: 1.49, 95% CI: 1.04- 2.15). Moreover, the age of the participants and the fear of Covid-19 were significantly associated with preventive medication use. People aged 60 and above were most likely to take preventive medicines, and the odds of taking medicines for those who were very afraid of the pandemic was 2.48 times (95% CI: 1.36- 4.53) than that of participants who were not afraid at all (table 3).

**Table 3:** Results of multivariable logistic regression of the factors associated with the use of medicines and herbal food/products among the public as a preventive measure against Covid-19

| Variables | Use of medicine as a preventive measure against Covid-19 |  |  | Use of herbal food/product as a preventive measure against Covid-19 |  |  |
| --- | --- | --- | --- | --- | --- | --- |
|  | Adjusted Odds ratios | 95% Confidence interval | P- value | Adjusted Odds ratios | 95% Confidence interval | P- value |
| **Gender** |  |  | 0.03 |  |  | 0.016 |
| Female | ref |  |  | ref |  |  |
| Male | 1.49 | 1.04- 2.15 |  | 0.73 | 0.57-0.94 |  |
| **Age** |  |  | 0.004 |  |  | 0.049 |
| 18-29 | ref |  |  | ref |  |  |
| 30-44 | 2.0 | 1.22- 3.27 |  | 0.62 | 0.43-0.90 |  |
| 45-59 | 1.02 | 0.49- 2.15 |  | 0.57 | 0.34-0.94 |  |
| 60 and greater | 2.93 | 1.37- 6.30 |  | 0.76 | 0.4-1.44 |  |
| **Residence** |  |  | 0.117 |  |  | 0.329 |
| Rural | ref |  |  | ref |  |  |
| Urban | 1.36 | 0.93- 2.0 |  | 0.87 | 0.66-1.15 |  |
| **Education** |  |  | 0.111 |  |  | 0.007 |
| Secondary or lower | ref |  |  | ref |  |  |
| Higher secondary | 1.43 | 0.84- 2.45 |  | 0.91 | 0.62-1.32 |  |
| Undergraduate | 1.89 | 1.13- 3.16 |  | 0.82 | 0.57-1.19 |  |
| Graduate | 1.38 | 0.79- 2.41 |  | 1.56 | 1.03-2.34 |  |
| **Marital status** |  |  | 0.238 |  |  | 0.034 |
| Unmarried | ref |  |  | ref |  |  |
| Married | 1.32 | 0.83- 2.11 |  | 1.44 | 1.03-2.02 |  |
| **Presence of chronic disease** |  |  | 0.776 |  |  | 0.682 |
| No | ref |  |  | ref |  |  |
| Yes | 1.07 | 0.66- 1.73 |  | 1.08 | 0.75-1.56 |  |
| **Fear of Covid-19** |  |  | 0.012 |  |  | <0.001 |
| Not afraid | ref |  |  | ref |  |  |
| Somewhat/<br>Moderately afraid | 1.89 | 1.07- 3.35 |  | 2.58 | 1.84-3.63 |  |
| Very afraid | 2.48 | 1.36- 4.53 |  | 2.03 | 1.39- 2.97 |  |

ref, reference

**Figure 2:**
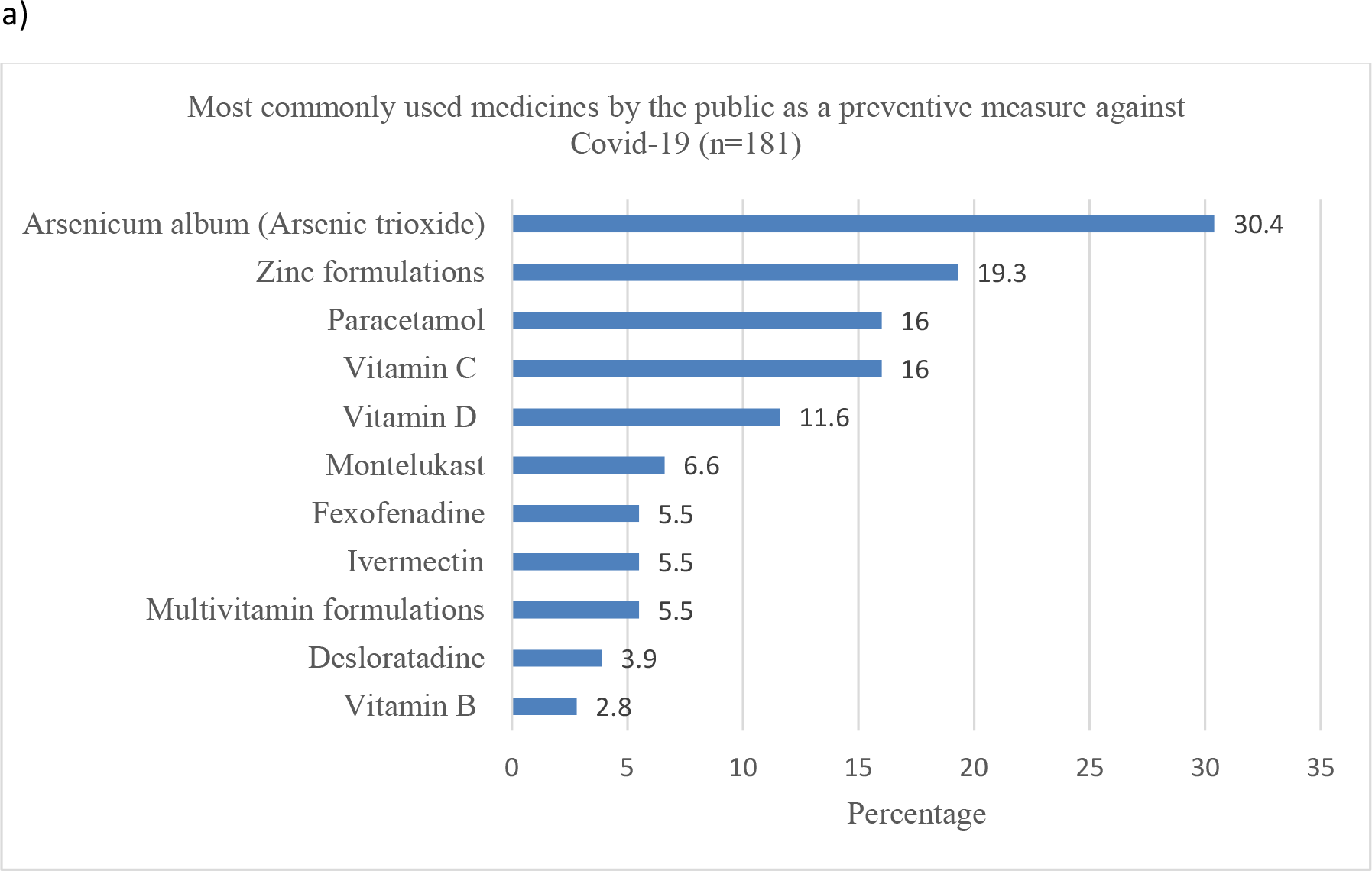
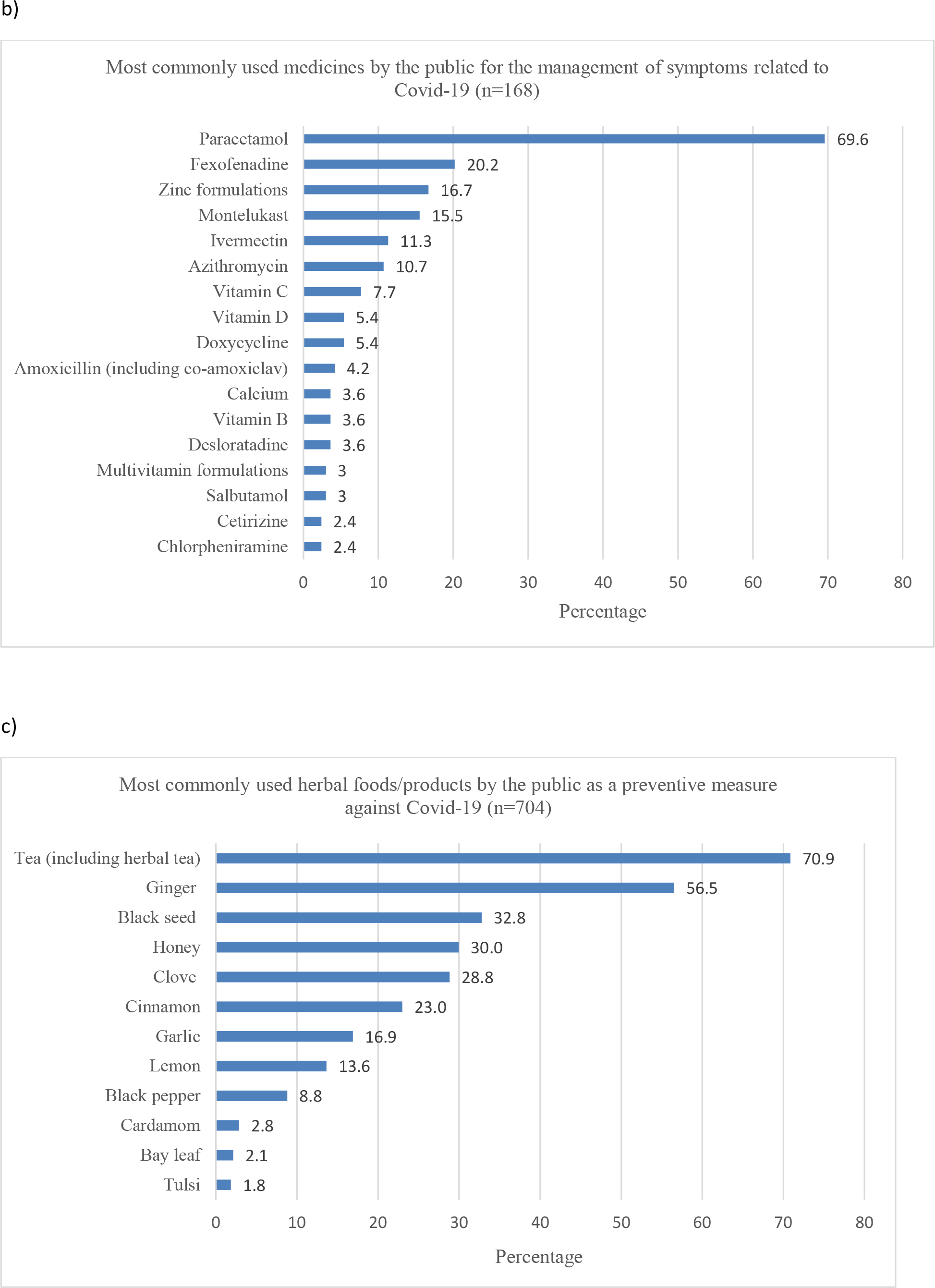
Use of medicine for prevention (a) and management (b) of Covid-19 symptoms and the use of herbal foods/products for prevention (c) of Covid-19 (the cumulative percentage may be greater than 100 since many participants took more than one drug or herb)

Around 20% (239) experienced one or more symptoms related to Coronavirus infections, and 70% (168) of them took medications to manage the symptoms. Paracetamol was the most commonly used drug, followed by Fexofenadine, Zinc, Montelukast, Ivermectin, etc. (Figure 2b). Also, 34 took antibiotics including Azithromycin, Doxycycline, Amoxicillin (including amoxicillin-clavulanic acid combination), Levofloxacin, Linezolid, and Cephalosporins.

### Use of herbal foods/products

A large number of participants (57.6%) reported having taken herbal foods/products to lower the risk of Covid-19 infection. About 71% of them took tea (normal and herbal), while other herbal foods such as ginger, black seed, honey clove, etc. were used by 56.5%, 32.8%, 30%, and 28.8% respectively (figure 2c). These herbs were often taken alone, or combined with tea or hot water. Factors significantly associated with taking herbal foods/products were being female, young (18–29 years), married, and afraid of the pandemic. Also, participants with a graduate degree were more likely to take preventive herbal foods/products than those with a secondary degree or lower (OR: 1.56, 95% CI: 1.03-2.34) (table 3).

### Source of information related to the use of medicines and herbal foods/products

When asked about the source of information related to their use of medicines or herbal foods/products, most participants said they had relied on the advice of family, friends, or relatives (40.1%) and their own knowledge (32.4%). Media (print, digital, and social) were also a major source of information. However, only 25% of the participants sought advice from Doctors while 18% from Pharmacists. (figure 3).

**Figure 3:**
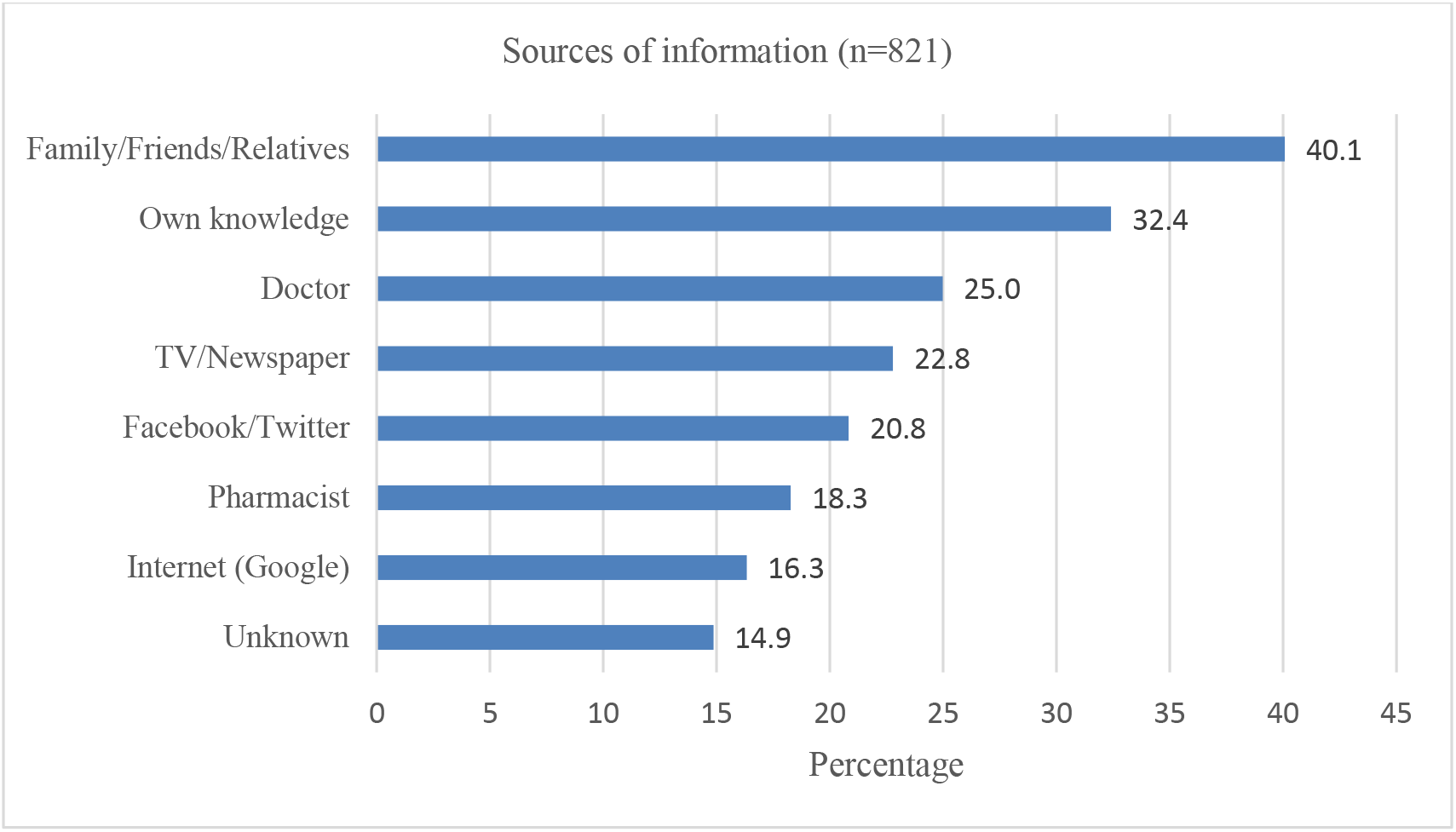
Sources of information related to the use of medicine and herbal food/product by the public (the cumulative percentage is greater than 100 since many participants relied on more than one source of information)

## Discussion

The present study was conducted at a time when the Covid-19 pandemic had reached its peak in Bangladesh [1], and we observed high rates of adoption of behavioral preventive measures (washing hands, staying home, avoiding crowds, and wearing masks), which was quite similar to the earlier studies conducted in Bangladesh [12–14]. Also, there was a decrease in cigarette smoking among half of the smokers. Studies have shown that smokers generally have a high risk of respiratory tract infections [15], and smoking can also be a risk factor for Covid-19 progression [16]. However, some participants reported an increase in smoking, which can probably be attributed to the heightened stress, fear, and boredom associated with the pandemic and the lockdown.

A significant percentage of the participants have taken prophylactic medications, and we have also identified potentially unnecessary and inappropriate use of certain medicines. For instance, the most used prophylactic medicine was the homeopathic medicine Arsenicum album. Arsenicum album has been recommended as a prophylactic drug against Covid-19 by Vellingiri et al. [17], but there is no clinical evidence supporting its effectiveness against Covid-19 [18]. While there are claims that Arsenicum album provides temporary relief against flu-symptoms, such claims are not proven and the United States Food and Drug Administration does not approve this drug [19]. Furthermore, homeopathy in general faces lots of criticism for being implausible, unscientific, and unreliable [20, 21]. Although it is hard to tell why so many people used this drug, one of the reasons may be the fact that the Ministry of Ayurveda, Yoga & Naturopathy, Unani, Siddha and Homoeopathy (AYUSH) of the neighboring country India recommended Arsenicum album for prophylactic use against Covid-19 [18]. The media may also play a role in the popularity of Arsenicum album. Moreover, many participants used Paracetamols prophylactically although it should only be used to manage symptoms associated with Covid-19. Another notable case was the prophylactic use of Ivermectin; out of the ten participants who used Ivermectin, only two sought advice from Doctors. Similar cases of Ivermectin self-medication has been reported in Brazil [22]. Prophylactic use of Ivermectin for Covid-19 is not only unproven, but it can also be quite risky, especially when self-medicated [22].

Since there is currently no effective treatment of Covid-19, the intake of vitamins, minerals, and herbs can serve to boost the immune system which can subsequently lower risks of infection and disease progression [7–9]. In this study, many participants reported having taken vitamin D, C, B and Zinc supplements. The use of herbal foods/products was also very high, probably because these are always available in most homes in Bangladesh and are known for their medicinal values. Green tea and black tea polyphenols have been reported to show antiviral activities and may have applications in Covid-19 prophylaxis and treatment [23]. Other herbal products such as ginger, garlic, honey, and black seed have also been mentioned for their antiviral actions and potential action against Covid-19 [24–26]. However, it should be mentioned that some of the reported cases of herbal foods/product usage in this study could be habitual and participants might have taken them without being aware of their association with Covid-19 prevention (for example tea).

When investigating the drugs used for management of symptoms related to Covid-19, our goal was to get an overview of the commonly used medicines in the community; assessment of hospital-based treatments or management of critical patients was beyond the scope of this study. Most people infected with Covid-19 experience only mild symptoms and intake of antipyretics (Paracetamol) may be sufficient for that, according to WHO [27]. The national guidelines on clinical management of Coronavirus disease 2019 recommend Paracetamol and Fexofenadine for the management of mild symptoms and suggest the intake of Zinc and Vitamin C [28]. Similar to the guidelines, in this study, Paracetamol, Fexofenadine, and Zinc have been found to be the most commonly used medicines for symptom management.

In the present study, fear of Covid-19 has been found to be the most important factor associated with preventive measures among the public; those who reported being afraid of Covid-19 adopted a greater number of preventive practices and were more likely to take preventive medicines and herbs. No other study has yet investigated the association between Covid-19 fear and medication use among the public, but in a study conducted in Turkey, fear of the pandemic has been found to be associated with a higher level of engagement to preventive practices [29]. Another study among the Thai healthcare workers found that those who reported fear and anxiety were more likely to adopt preventive practices such as washing hands and wearing masks and personal protective equipment [30].

Since the beginning of the pandemic, television, and social media (Facebook, Twitter) have been playing a big role in the spread of misinformation surrounding Covid-19 [31, 32]. We have found a high reliance of the participants on non-medical sources for medication-related advice pertaining to Covid-19, and so it is important that the spread of misinformation through digital, print, and social media is contained as much as possible.

## Conclusions

In summary, we have observed a high adoption of preventive measures by the public. Also, a considerable number of participants took preventive medicines and herbal products. Fear of Covid-19 has been identified as one of the most important factors associated with the adoption of behavioral preventive measures as well as the use of preventive medicines and herbal foods/products. For information pertaining to the use of medicines and herbs, most participants relied on non-medical sources. Moreover, potential misuse and unnecessary use of certain drugs have been identified, most probably due to the fear and panic surrounding Covid-19, the spread of misinformation through the media, and restricted access to healthcare providers during the lockdown.

## Data Availability

All data collected, without personally identifiable information, is available upon request to the corresponding author

## Ethical approval

The research protocol was reviewed and approved by the Antimicrobial Resistance Action Center (ARAC), Animal Health Research Division, Bangladesh Livestock Research Institute (BLRI), Bangladesh (Approval no: ARAC:15/06/2020:03)

